# Reproduction numbers for epidemics with hidden cases, re-infections and newborns

**DOI:** 10.1101/2025.05.28.25328507

**Authors:** Igor Nesteruk

## Abstract

Real-time assessments of the reproduction rates are crucial for timely responses to changes in epidemic dynamics. Known effective reproduction numbers *R*_*t*_ are based on registered (visible) cases despite that asymptomatic and unregistered patients occur in all epidemics and need to be corrected to take into account the number of hidden cases. Since newborns and re-infections significantly affect the dynamics of epidemics, they should also be taken into account in calculations of *R*_*t*_ and recently proposed reproduction rates *R*_*τ*_ - the ratios of the real numbers of infectious persons at different moments of time. The numbers of cases generated by symptomatic and asymptomatic patients were introduced, estimated using a novel mathematical model and compared with the results of classical SIR (Susceptible-Infectious-Removed) model for the COVID-19 pandemic dynamics in Austria. Reproduction rates *R*_*τ*_ were estimated with the use of visible accumulated numbers of COVID-19 cases in Austria and Tanzania (including real-time approach). The proposed methods for calculating reproduction numbers may better reflect the COVID-19 pandemic dynamics than the results listed by John Hopkins University.

## 1. Introduction

Effective reproduction numbers *R*_*t*_ [1–9] are based on the registered (visible) cases *V*^*(v)*^. Since the real numbers of cases *V* can be higher due to asymptomatic and unregistered patients [10–17], new reproduction numbers must be introduced taking into account also the influence of re-infections [18–21] and newborns [22, 23]. In particular, new reproduction numbers - the ratios of the real numbers of infectious persons at different moments of time – were proposed in [24].

In this study we will generalize the known effective reproduction numbers *R*_*t*_ in order to take into account the unregistered cases and discuss the methods of their estimations with the use of novel mathematical model [23]. Numerical results for the COVID-19 pandemic in Austria will be presented. The numbers of cases generated by symptomatic and asymptomatic patients will be estimated and compared with the results of classical SIR (Susceptible-Infectious-Removed) model [8,9] and calculations of the effective reproduction number *R*_*t*_ based on the method proposed in [5] and listed in [25].

New reproduction numbers *R*_*τ*_ (introduced in [24]) will be estimated with the use of visible accumulated numbers of COVID-19 cases in Austria and Tanzania listed in [25]. Corresponding values will be compared with the effective reproduction numbers *R*_*t*_ reported in [25] and calculated using recommendations of the Robert Koch Institute (RKI) [11].

Real-time assessments of the reproduction rates are crucial for timely responses to changes in epidemic dynamics. The new methods for real-time calculations of *R*_*τ*_ will be proposed and the results of calculations will be compared with the daily numbers of COVID-19 cases registered in Austria in 2020.

## 2. Materials and Methods

### 2.1. Effective Reproduction Numbers for the Visible and Hidden Epidemic Dynamics

The effective reproduction number *R*_*t*_*(t)* shows the average number of people infected by one person [1, 2] versus of time *t*. Since these infections can be visible (registered) with accumulated numbers of cases *V*^*(v)*^ and asymptomatic (hidden and unregistered, *V*^*(h)*^), we will use the novel mathematical model (1)- (5), [23]

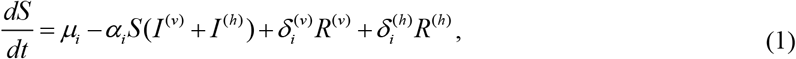

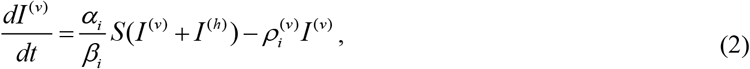

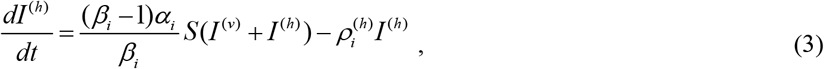

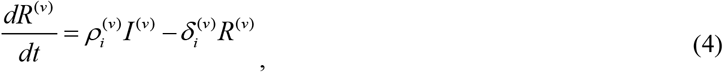

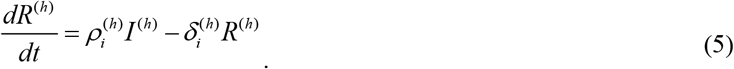

with the compartment of infectious persons *I(t)* divided into visible (registered) *I* ^(*v*)^ and hidden *I* ^(*h*)^ (invisible/asymptomatic and unregistered) parts (*I* = *I* ^(*v*)^ + *I* ^(*h*)^, see eqs. (1)-(5)). The visibility coefficient *β*_*i*_ ≥ 1 is the ratio of total infections to the visible ones (*β*_*i*_ = 1 corresponds to the fully visible epidemic). The compartment of removed persons *R(t)* is also divided into visible (registered) and hidden parts *R* = *R*^(*v*)^ + *R*^(*h*)^ with removing rates 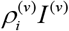 and 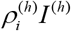, respectively (see eqs. (4) and (5)). We suppose the waning immunity rates to be proportional to the number of removed persons 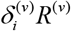 and 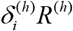, [23]. Corresponding terms appear in eqs. (4) and (5) with a minus sing and in eq. (1) with a plus sign. Due to newborns, the number of susceptible persons *S(t)* increases at constant rate *μ*_*i*_ (see eq. (1)). Infection, removal and waning immunity rates 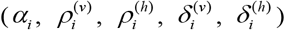, the visibility coefficient *β*_*i*_ and the increasing rate of the susceptible persons *μ*_*i*_ are supposed to be constant for every epidemic wave, i.e. for the time periods: 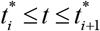, *i* =1, 2,3,….

Let us introduce two effective reproduction numbers 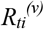 and 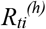 showing the numbers of visible and hidden patients, respectively, infected by one person (symptomatic or asymptomatic). With the use of total numbers of infectious *I* = *I* ^(*v*)^ + *I* ^(*h*)^ (the sum of visible and hidden ones) and average durations of spreading the infection 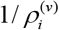 and 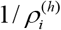 for visible and asymptomatic patients during the *i-th* epidemic wave (corresponding removal rates 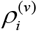 and 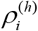 can be different, see eqs. (1), (4) and (5)), the following formulae can be obtained :

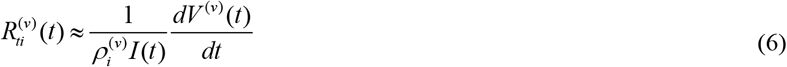

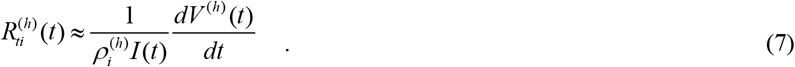

Since new infections (derivatives in eq. (6) and (7)) are appearing according to the visibility coefficient *β*_*i*_≥ 1 and the infection rate *α* :

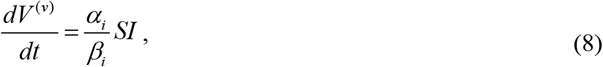

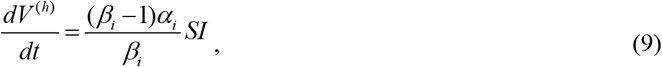

eqs. (6)–(9) yield:

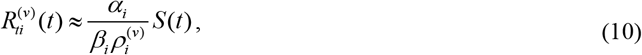

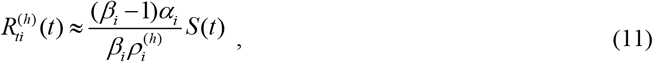

For the fully visible epidemic (*β* = 1), eq. (10) coincide with one obtained in [8] and is in good agreement (see [9]) with the Robert Koch Institute (RKI) recommendations to calculate the reproduction number for the COVID-19 pandemic as “the ratio of new infections in two consecutive time periods each consisting of 4 days”, [11]. In terms of the smoothed accumulated numbers of visible cases [8, 27]

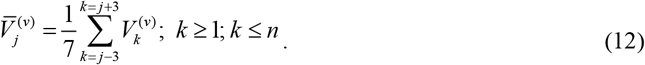

the RKI rule can be written as follows, [9]:

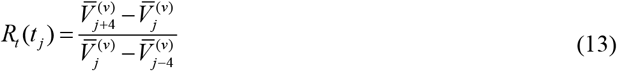

The effective reproduction number *R*_*ti*_ representing the total number of patients (registered and hidden) infected by one person is the sum of (10) and (11):

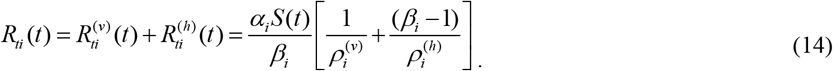

If the removal rates for symptomatic and asymptomatic patients are equal 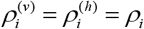, eq. (14) does not depend on the visibility coefficient and coincide with one obtained in [8]:

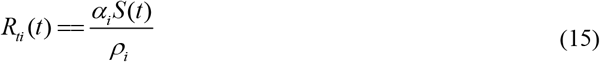

### 2.2. Reproduction Numbers *R*_*τ*_(*t*) for Epidemics with Re-infections and Newborns

A new reproduction number

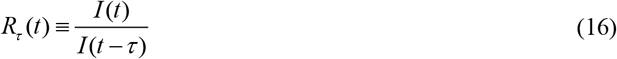

was proposed in [24] to indicate the rate of increasing (decreasing when *R*_*τ*_(*t*) <1) the real number of infectious persons (symptomatic and asymptomatic) during time *τ* and estimated for the case 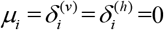. To take into account newborns and re-infections, let us use relationship (8) in eq. (16). Then

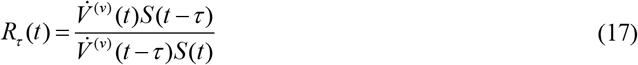

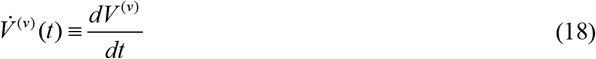

Reproduction numbers in formulae (10), (11), (14)–(17) can be calculated by a numerical integration of set (1)-(5) using initial conditions listed in [23]. To identify the values of 13 unknown parameters, the least squares method can be used [23, 26]. Usually, the number of susceptible persons *S* is very large [8, 14–16, 23, 27] and *S*(*t* −*τ*) / *S*(*t*) ≈ 1for small enough values of *τ*. Then eq. (17) yields

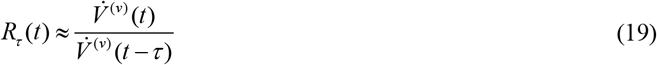

Formula (19) demonstrates that the new reproduction number *R*_*τ*_(*t*) does not depend on the model parameters and can be approximately estimated with the use of daily, weekly or monthly (according to the unit of time) numbers of new visible cases. In particular, the following formula can be used, [28]:

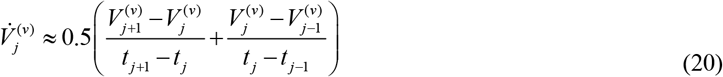

where 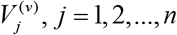 are the accumulated numbers of visible cases registered at moments *t*_*j*_. Parabolic interpolation yields the following estimation of daily, weekly or monthly numbers of new visible cases number of cases:

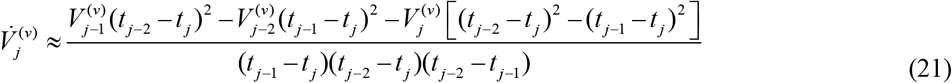

which does not contain unknown results of future observation and can be used instead (20) for real-time estimations. Usually, daily characteristics are very random (e.g., for the COVID-19 pandemic, [25]), show some weekly periodicity and need smoothing [8, 9, 24, 25, 27]. In particular, the smoothed accumulated numbers of cases (12) can be used.

Real-time estimations of reproduction numbers (17) at moment *t* = *t_j_* do not allow using unknown results of future observations 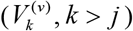 and eqs. (12), (13), and (20). To estimate derivatives in formula (19), smoothed daily numbers of new cases, calculated with the use of previous observations only, can be applied. Since the observations obtained closer to the moment *t* = *t_j_* have stronger impact on the trends, some weighing coefficients and following values:

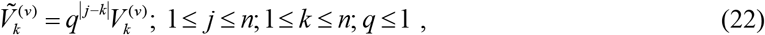

can be used in 7-days smoothing:

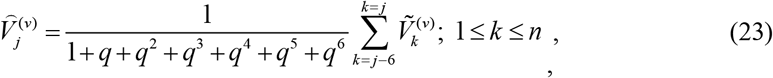

The value of parameter *q* has to be adjusted for every particular case. For example, the value *q*=1 was used in John Hopkins University (JHU) datasets to calculate the smoothed numbers of daily COVID-19 cases [25].

## 3. Results and Discussions

### 3.1. Calculations of Effective Reproduction Numbers for the COVID-19 Pandemic in Austria

Fig. 1 represents JHU datasets [25] (version corresponding to 11 December 2023) for number of accumulated (*AC,* blue “crosses”) and daily (*DC*, smoothed, black “crosses”) registered in Austria in 2020 and results of calculations of the effective reproduction number *R*_*t*_*(t)* (according to [5], for fully visible epidemic, red “crosses”). To investigate the influence of hidden cases, the set of differential equations (1)-(5) were integrated numerically with the use of initial conditions listed in [23] and the fourth order Runge-Kutta method [29]. No method of least squares was applied to detect the optimal values of parameters. We have used only parameter sets that provide some similarity to the observed *AC* and *DC* figures registered in April 2020. The solid black curve corresponds to the visible (registered) daily numbers of COVID-19 cases (eq. (8)); the solid blue curve represent the theoretical number of accumulated cases (integral of (8)) for the following values of parameters: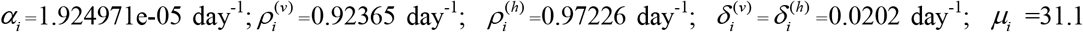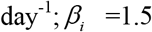. The green, magenta and solid red curves show the effective reproduction numbers (visible, eq. (10); hidden, eq. (11) and total, eq. (14)).

**Fig. 1.**
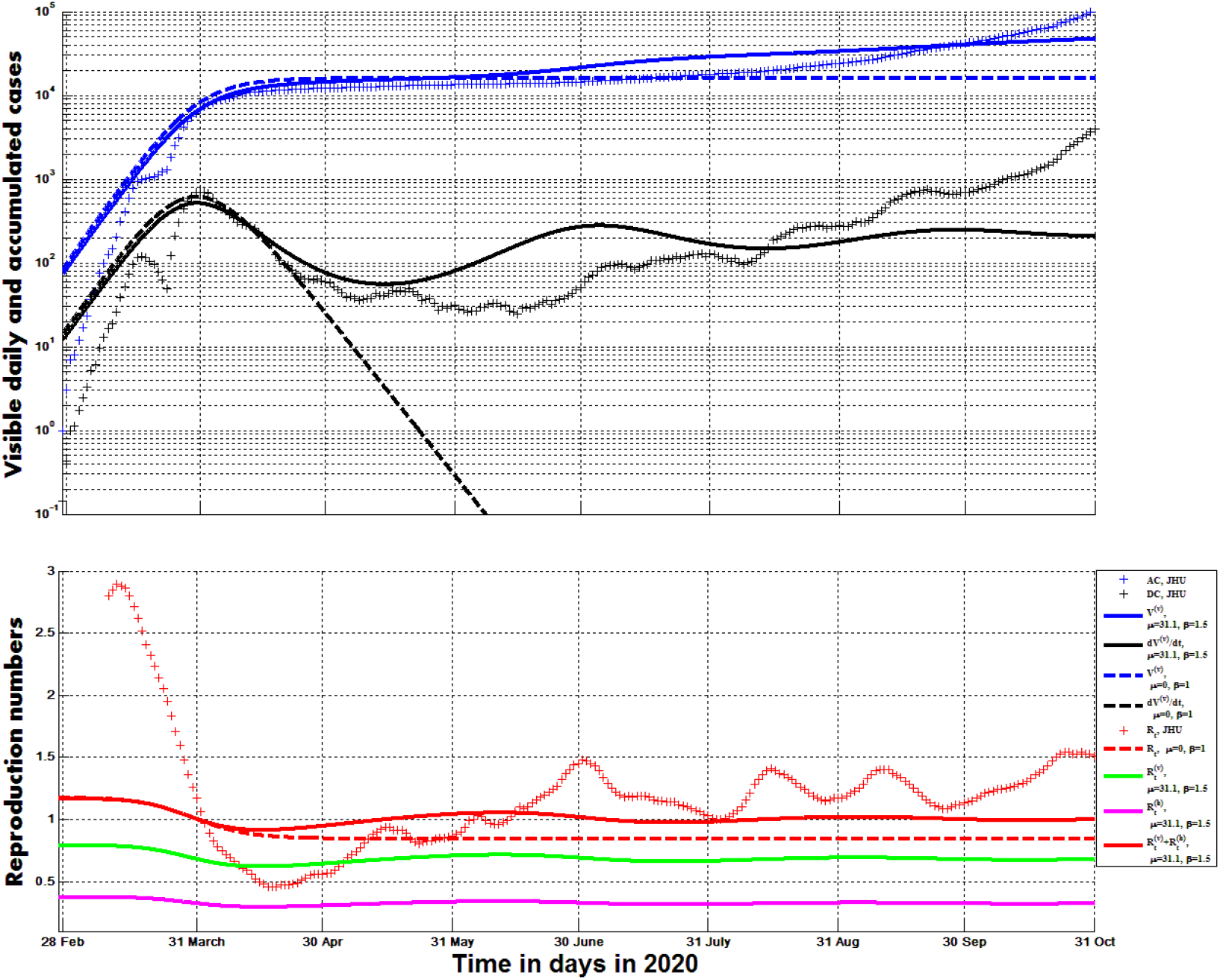
The COVID-19 pandemic dynamics in Austria in 2020. Registered daily and accumulated numbers of cases and calculations of the reproduction numbers. Blue and black “crosses” represent visible (registered) accumulated numbers of cases *AC* and smoothed daily numbers of new cases *DC*, respectively, [25]. Red crosses correspond to the calculations of the effective reproduction number by the method proposed in [5] listed in [25]. Curves show the results of numerical integration of (1)-(5) at different values of parameters: blue – visible accumulated number of cases *V* ^(*v*)^; black – visible daily number of cases *dV* ^(*v*)^ / *dt*; magenta - the hidden reproduction number 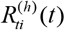, eq. (11); green – the visible reproduction number 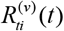, eq. (10); red - the total reproduction number 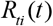, eq. (14).

The dashed curves in Fig. 1 correspond to the fully visible epidemic with 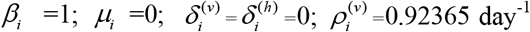, which can be simulated with the use of the classical SIR model [8, 22, 27, 30–33]. Black line yields the theoretical estimation of the daily numbers of cases (eq. (8)); blue – accumulated number of cases; red – reproduction number (10). According to the classical SIR model, *S(t)* values and the reproduction number decrease monotonously with time (see eq. (1) at 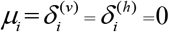, eq. (10) and the dashed red curve in Fig. 1). When the re-infections and newborns are taken into account, the oscillations in reproduction numbers are possible (see solid magenta, green and red curves) In particular, the total reproduction number (eq. (14) and the red solid line) several times reached the critical value 1.0. The method proposed in [5] (see red “crosses” in Fig.1) yields much higher magnitude of oscillations, but the moments of time, corresponding to the critical values of the reproduction number were very close until mid-June 2020 (compare with the solid red line). Probably the coincidence can be increased with the use of optimal parameter sets corresponding to the later epidemic periods in (1)-(5).

### 3.2. Comparisons of Different Reproduction Rates and Methods of Their Calculations

In Fig. 2 the results of calculations of the effective reproduction number *R*_*t*_*(t)* (according to [5], for fully visible COVID-19 epidemic in Austria, red “crosses”, JHU dataset [25]) are compared with the RKI method (eqs. (12) and (13), the magenta curve). The values shown by red crosses ([5, 25]) diminished almost monotonously until mid-April 2020 and did not reflect local minimum and huge increase in the smoothed daily numbers of new cases *DC* (calculated by using the JHU dataset in eq. (12) and substituting into formula (20), the black curve). In comparison, the RKI method and calculations of *R*_*τ*_(*t*) (blue and brown curves) reflect these changes in epidemic dynamics yielding huge increase of reproduction rates. The *R*_*τ*_(*t*) values (19) at *τ* =1 day calculated by putting smoothed values (12) into (20) (the blue curve) start to increase simultaneously with *DC*. When the smoothed *DC* values listed by JHU are used in (19) (the brown curve), some delay occurs (compare with black and blue lines).

**Fig. 2.**
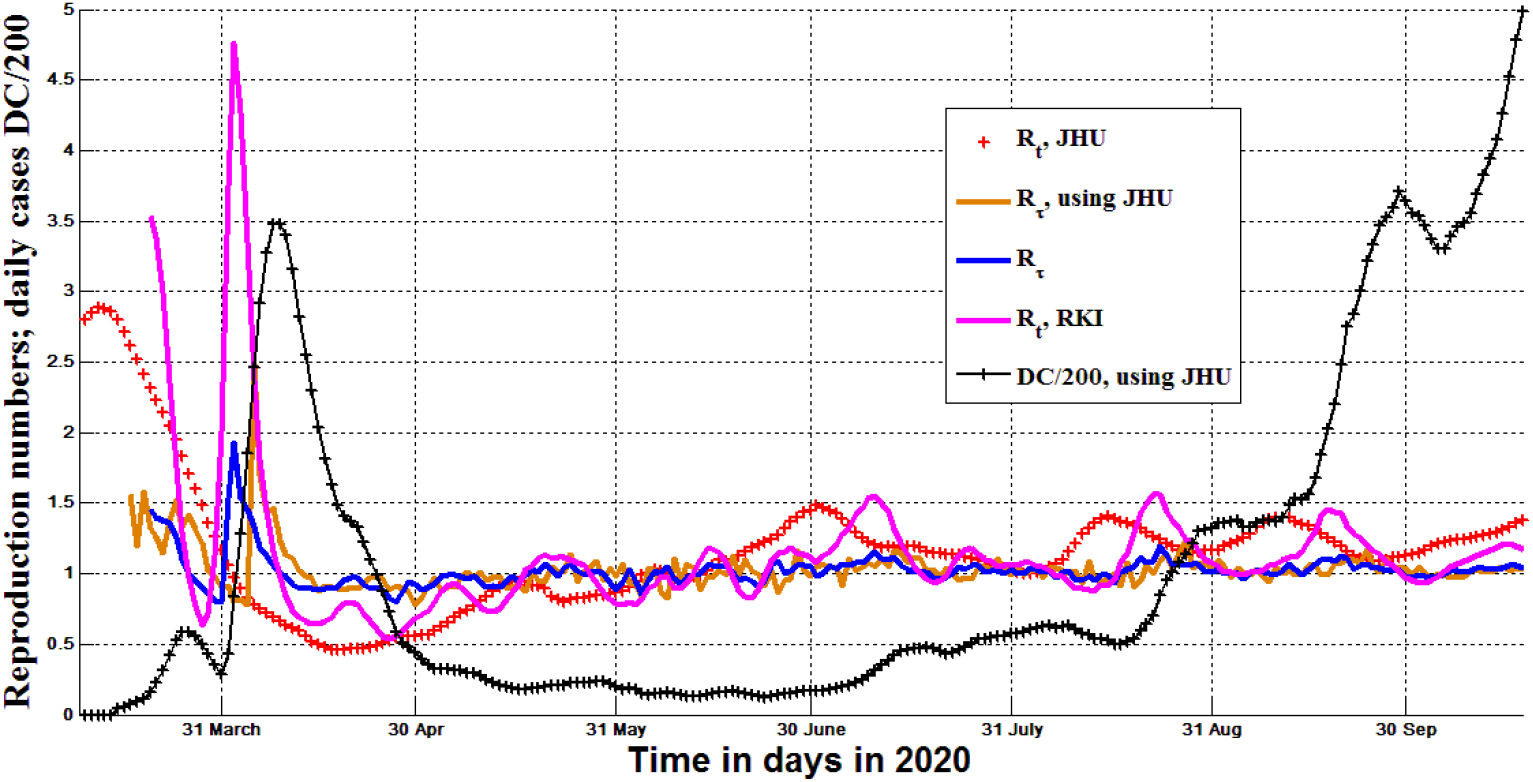
Reproduction rates and the smoothed daily number of COVID-19 cases in Austria in 2020. The black curve represents visible (registered) daily number of cases *DC* smoothed by using the accumulated numbers 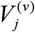 listed in JHU dataset [25], eqs. (12) and (20). Red “crosses” correspond to the calculations of the effective reproduction number *R*_*t*_ by the real-time method proposed in [5] (the results are listed in [25]). The magenta line represents *R*_*t*_ values calculated by the RKI method (eqs. (12), (19) and (20)). Blue and brown curves show calculations of *R*_*τ*_(*t*) (eq. (19), *τ* =1 day). The brown one was obtained using the smoothed *DC* values listed by JHU [25]; blue - by putting smoothed values (12) into (20).

In Fig. 3, the results of calculations of the reproduction rates *R*_*t*_*(t)* and *R*_*τ*_(*t*) are compared with the COVID-19 pandemic dynamics in Tanzania (the accumulated number of cases *AC* is shown by blue “triangles”). Red “crosses” correspond to the calculations of the effective reproduction number *R*_*t*_ by the real-time method proposed in [5] (the figures are listed in [25]). These values diminished monotonously and did not reflect the huge increase in the numbers of cases registered in April 2020 (in comparison with the *R*_*t*_ values calculated by RKI method (the magenta curve)). It must be noted that all *R*_*t*_ figures listed by JHU [25] for Tanzania are subcritical (*R*_*t*_ <1), while for an epidemic outbreak (or a new wave) to occur, we need to have *R*_*t*_ >1. Blue and black curves show calculations of *R*_*τ*_(*t*) (eq. (19) at *τ* =1 day). The black one was obtained using the smoothed *DC* values listed by JHU [25]; blue - by putting smoothed values (12) into (20). All curves in Fig.3 show some supercritical values and reflect the increase in the number of cases. The black line is more chaotic and reflects the increase in the number of cases with some delay (in comparison with the blue one).

**Fig. 3.**
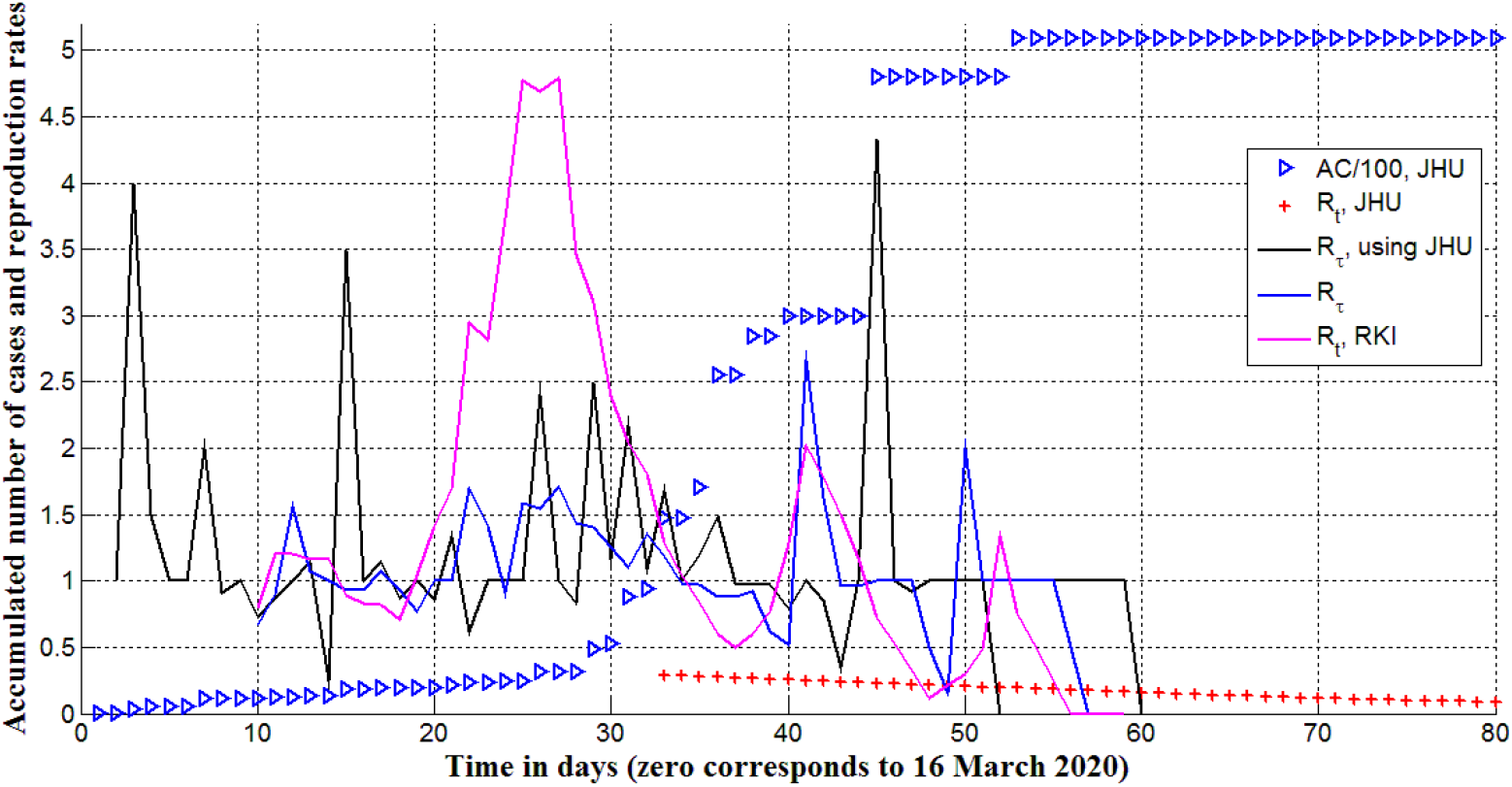
Reproduction rates and the accumulated number of COVID-19 cases in Tanzania in 2020. Blue “triangles” represent visible (registered) accumulated number of cases *AC* divided by 100 (according to the JHU dataset [25]). Red “crosses” correspond to the calculations of the effective reproduction number *R*_*t*_ by the method proposed in [5] and listed in [25]. The magenta line represents *R*_*t*_ values calculated by the RKI method (eqs. (12), (19) and (20)). Blue and black curves show calculations of *R*_*τ*_(*t*) (eq. (19), *τ* =1 day). The black one was obtained using the smoothed *DC* values listed by JHU [25]; blue - by putting smoothed values (12) into (20).

Thus, some estimations of *R*_*t*_ cannot reflect changes in epidemic dynamics (compare red “crosses” with *DC* and *AC* values in Figs. 2 and 3). In comparison, the presented calculations of *R*_*τ*_(*t*) demonstrated increase and supercritical values for the periods, when the daily number of cases increased. Unfortunately, the real-time estimations of *R*_*τ*_(*t*) show some time delay (compare the brown and black curves with the blue one in Figs. 2 and 3). This fact limits the applicability of the real-time approaches for effective control of the epidemic dynamics. Nevertheless, the delay can be diminished with the use of weighing coefficients (see eq. (22) and (23)).

In Fig. 4 different methods of calculations of the reproduction rate *R*_*τ*_(*t*) are presented and compared with the daily numbers of COVID-19 cases in Austria smoothed with the use of AC values listed in [25], eqs. (12) and (20) and divided by 500. Other curves show calculations of *R*_*τ*_(*t*) (eq. (19) at *τ* =1 day). Blue one was calculated by putting values (12) into (20) (median non real-time smoothing). Red, magenta and green curves correspond to the real-time estimations using smoothed *DC* values according to the JHU dataset [25] (red); obtained by putting smoothed values (23) into (21) at *q*=1 (magenta) and *q*=0.5 (green).

**Fig. 4.**
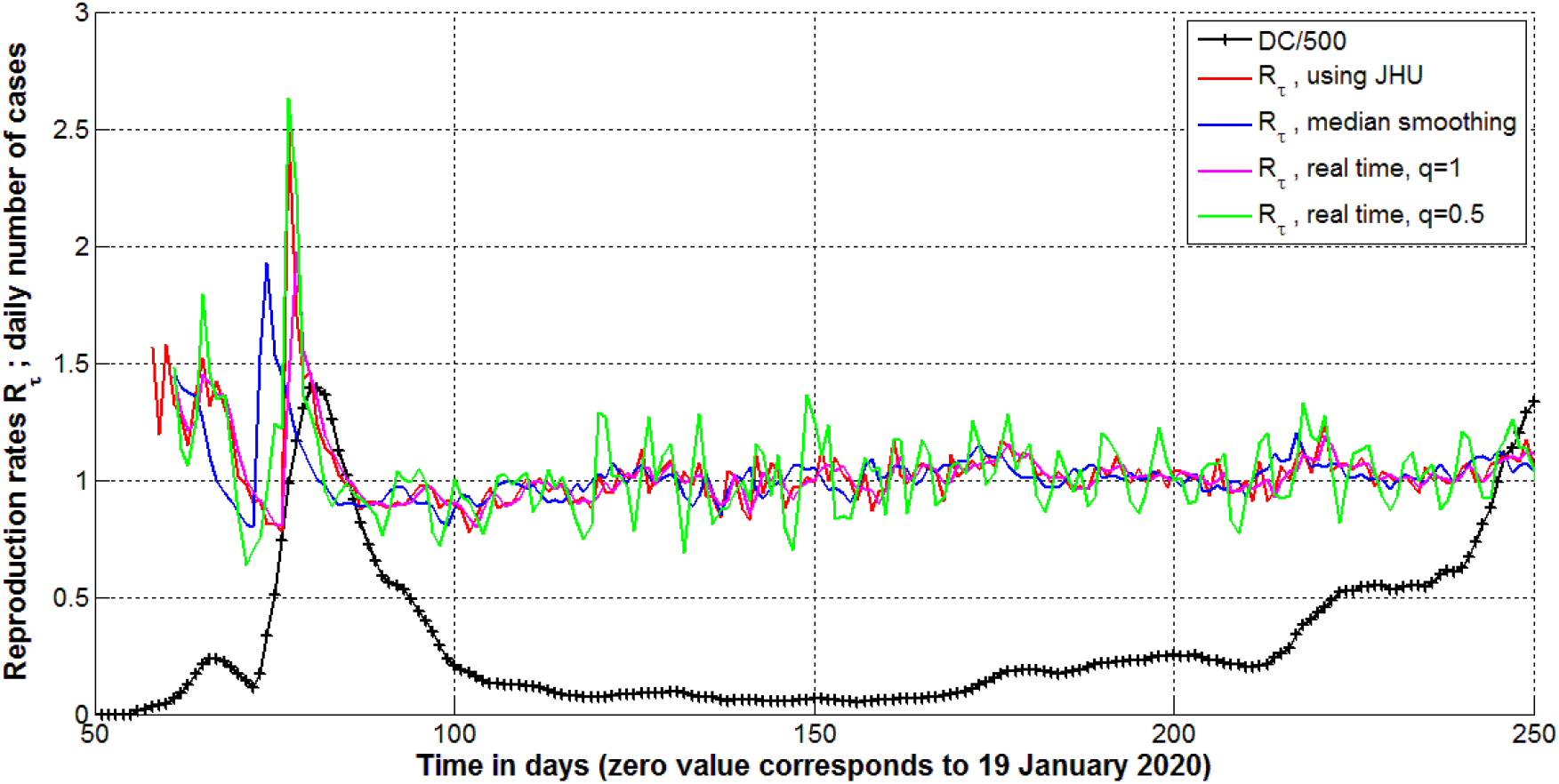
Reproduction rates *R*_*τ*_(*t*) and smoothed daily number of COVID-19 cases in Austria in 2020. The black curve represent visible (registered) daily number of cases *DC* smoothed using the accumulated numbers 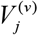 listed in JHU dataset [25], eqs. (12) and (20) and divided by 500. Other curves show calculations of *R*_*τ*_(*t*) (eq. (19), *τ* =1 day). Blue one was calculated by putting values (12) into (20) (median non real-time smoothing). Red, magenta and green curves correspond to the real-time estimations using smoothed *DC* values according to the JHU dataset [25] (red); obtained by putting smoothed values (23) into (21) at *q*=1 (magenta) and *q*=0.5 (green).

Red and green curves are very close illustrating the fact that eq. (19) allows us to calculate reproduction rates *R*_*τ*_(*t*) using smoothed *DC* values for fixed and previous moments of time or eqs. (21)–(23) at *q*=1. The value *q*=0.5 yields more chaotic behavior of *R*_*τ*_(*t*) (green curve) but decreases the delay between changes in *R*_*τ*_(*t*) and the epidemic dynamics. Probably, the value of *q* can be optimized for every epidemic. For some diseases only monthly or weekly numbers of new cases are available (e.g., the pertussis epidemic in England, [34]). Then there is no need to use smoothing and real-time estimations of *R*_*τ*_(*t*) can be obtained using (19) and (21).

## 4. Conclusions

The impact of newborns and re-infections on the visible and hidden epidemic dynamics and reproduction rates was taken into account with the use of a novel mathematical model. The known reproduction rate *R*_*t*_ was generalized to show the numbers of cases generated by symptomatic and asymptomatic patients, which were calculated for the COVID-19 pandemic in Austria in 2020. When the re-infections and newborns are taken into account, the oscillations in reproduction numbers are possible. In particular, the total reproduction number (the sum of number of cases generated by symptomatic and asymptomatic patients) several times reached the critical value 1.0.

Methods for calculations of the recently proposed reproduction rates *R*_*τ*_ - the ratios of the real numbers of infectious persons at different moments of time – are presented and estimated with the use of visible accumulated numbers of COVID-19 cases in Austria and Tanzania (including real-time approach). The proposed methods for calculating reproduction numbers may better reflect the COVID-19 pandemic dynamics than the results listed by John Hopkins University.

Unfortunately, the real-time estimations of *R*_*τ*_(*t*) show some time delay. This fact limits the applicability of the real-time approaches for effective control of the epidemic dynamics. Nevertheless, the delay can be diminished with the use of weighing coefficients. The search for optimal values of these coefficients may be a topic for further research.

## Clarification point

No humans or human data was used during this study

## Data Availability

All data produced in the present work are contained in the manuscript

## Data availability

Only open datasets available on Internet were used in this study.

## Acknowledgements

The author is grateful to Ulrike Tillmann, James Robinson, Robin Thompson, Matt Keeling, Paul Brown, and Oleksii Rodionov for their support and providing very useful information. This paper was written with the support of the INI-LMS Solidarity Programme at the University of Warwick, UK.

